# Cost & Cost-Effectiveness of Implementing SD Biosensor Antigen Detecting SARs-CoV-2 Rapid Diagnostic Tests in Kenya

**DOI:** 10.1101/2023.01.05.23284225

**Authors:** Brian Arwah, Samuel Mbugua, Jane Ngure, Mark Makau, Peter Mwaura, David Kamau, Desire Aime Nshimirimana, Edwine Barasa, Jesse Gitaka

## Abstract

The COVID-19 pandemic has created a need to rapidly scale-up testing services. In Kenya, services for SARS-CoV-2 nucleic acid amplifying test (NAAT) have often been unavailable or delayed, precluding the clinical utility of the results. The introduction of antigen-detecting rapid diagnostic tests (Ag-RDT) has had the potential to fill at least a portion of the ‘testing gap’. We, therefore, evaluated the cost-effectiveness of implementing SD Biosensor Antigen Detecting SARs-CoV-2 Rapid Diagnostic Tests in Kenya.

We conducted a cost and cost-effectiveness of implementing SD biosensor antigen-detecting SARS-CoV-2 rapid diagnostic test using a decision tree model following the Consolidated Health Economic Evaluation Standards (CHEERS) guidelines under two scenarios. In the first scenario, we compared the use of Ag-RDT as a first-line diagnostic followed by using NAAT assay, to the use of NAAT only. In the second scenario, we compared the use of Ag-RDT to clinical judgement. We used a societal perspective and a time horizon of patient care episodes. Cost and outcomes data were obtained from primary and secondary data. We used one-way and probabilistic sensitivity analysis to assess the robustness of the results.

At the point of care, Ag-RDT use for case management in settings with access to delayed confirmatory NAAT testing, the use of Ag-RDT was cost-effective (ICER = US$ 964.63 per DALY averted) when compared to Kenya’s cost-effectiveness threshold (US$ 1003.4). In a scenario with no access to NAAT, comparing the Ag-RDT diagnostic strategy with the no-test approach, the results showed that Ag-RDT was a cost-saving and optimal strategy (ICER = US$ 1490.33 per DALY averted).

At a higher prevalence level and resource-limited setting such as Kenya, implementing Ag-RDT to complement NAAT testing will be a cost-effective strategy in a scenario with delayed access to NAAT and a cost-saving strategy in a scenario with no access to NAAT assay.

## Introduction

The COVID-19 pandemic has created a need to rapidly scale-up testing services and provide diagnoses to implement test-trace-isolate strategies, essential to treat and care for patients and to control the spread of the virus. Hundreds of diagnostic products are now available on the market, targeting the detection of viral RNA, viral antigens, and host antibodies against SARS-CoV-2.

Services for SARS-CoV-2 Nucleic acid amplification testing (NAAT) assays have often been unavailable or backlogged for several days to weeks, precluding the clinical utility of the results. NAAT, a reverse transcription polymerase chain reaction (PCR) molecular testing of respiratory tract samples, is the recommended method for confirmation of COVID-19. In low and middle-income countries, however, the availability and health impact of PCR testing can be jeopardized by lack of testing capacity, insufficient trained personnel, shortages of reagents, long turnaround times (TAT), and high costs [1]. Lateral flow antigen-detecting rapid diagnostic tests (Ag-RDTs), which are easy to perform and provide results within 15-30 minutes, have recently been commercialized and have the potential to fill at least a portion of the ‘testing gap’. Under certain conditions, Ag-RDTs that meet minimum performance requirements are recommended, and some have WHO Emergency Use Listing authorization [2]. These simple-to-use tests offer the possibility of rapid case detection, especially of the most infectious patients in the first week of illness, at or near the point of care.

WHO released an interim guidance on the use of Ag-RDTs for SARS-CoV-2, and the use of Ag-RDTs is recommended when PCR is either unavailable or long TAT of PCR which delays its clinical utility. This is particularly the case in less privileged countries in Africa, especially in Sub-Saharan Africa [3].

National norms and policies are being adopted in Kenya and many countries to allow and encourage targeted use of these Ag-RDTs. The decision to fully implement rapid diagnostic kits for detecting SARS-CoV-2 in Kenya relies on the field performance, feasibility, acceptability, and cost-effectiveness of the RDT compared to other diagnostic methods in the different settings which involve point-of-care diagnosis of COVID-19.

A published study of Ricks et al. analyzed the health system cost and health impact of using RDTs among hospitalized and symptomatic patients with SARS-COV2, and confirmed that despite the low sensitivity of RDTs compared with RT-PCR, the Ag-RDTs have the potential to be more impactful with less cost per death and more infections averted [4]. Studies have focused on effectiveness of testing kits and leaving behind cost effectiveness research however, a specific approach to assess cost-effectiveness of health interventions suggested by the commission on macroeconomics and Health (WHO,2001) is that interventions costing less than the per capita gross domestic product (GDP) in Low and Middle-Income Countries (LMICs) are “very cost-effective”, and those costing less than triple the per capita GDP are “cost-effective [5]. This, therefore, raises one question, under which scenario in the point-of-care diagnosis is RDT cost-effective? The objective of this study was to evaluate the cost effectiveness of SD Biosensor Antigen Detecting SARs-CoV-2 Rapid Diagnostic Tests in Kenya.

## Methods

### Study Design

We developed a decision tree model in TreeAge Pro Healthcare 2021 to evaluate the cost-effectiveness of implementing SD biosensor antigen detecting SARS-CoV-2 rapid diagnostic tests in Kenya from a societal perspective. We modeled the costs and outcomes for diagnosing and treating COVID-19 patients in line with WHO interim guidance on antigen detection for COVID-19 using rapid immunoassays [6] and the Kenya ministry of health COVID-19 case management guidelines [7].

The diagnostic and treatment pathway followed the cases where symptomatic patients with suspected COVID-19 and asymptomatic contacts of COVID-19 cases attend health facilities with i) no access to NAAT for diagnosis or ii) limited access with prolonged turnaround times precluding clinical utility of results.

#### Evaluation Scenarios

We assessed two scenarios:

##### Point of care Ag-RDT use for case management in settings with access to delayed confirmatory NAAT testing scenario

The scenario represented a health facility that sends samples to an external lab for NAAT, often with delayed result reporting. In this situation, Ag-RDT would be the first-line test to allow for case detection and rapid implementation of isolation procedures amongst positives and prioritization of negatives for confirmatory testing by NAAT at a designated laboratory facility. We compare the scenario with patients subjected to NAAT test, which is associated with a long turn-around time but obviates the need for confirmatory testing of negatives or a case whereby there is no testing. The diagnosis only relies on the clinical presentation of COVID-19 symptoms as per WHO case definitions [8].

##### Point of care Ag-RDT use for case management in settings with no access to confirmatory NAAT testing scenario

The scenario differs from the first one since the target location involves a health facility with no access to NAAT and no secure means for the safe and timely transport of samples to centralized facilities. The scenario presents a case whereby Ag-RDT is the only feasible tool to aid in the diagnosis or a choice of not conducting a COVID-19 test.

### Sampling and sample size

We selected two counties in Kenya, Kiambu, and Nairobi counties, to assess the field performance, feasibility, acceptability, and impact of SD biosensor antigen detecting SARS-CoV-2 rapid diagnostic tests. The two counties were chosen since they had the highest prevalence of COVID-19 in Kenya (average at >3% over the past three months), they also had different levels of government-owned facilities, and they had communities that were at high risk of outbreaks. We sampled four facilities that captured the diversity of access for COVID-19 testing and drew a sample size of 18 patients to capture the patient cost perspective. The patients’ sample size was selected to achieve balance in the facilities chosen and we settled on the 18 patients after reaching saturation.

Using the previously proposed method by Buderer 1996, the initial sample size needed for the COVID-19 RDT assumed the following expected values: test sensitivity of 80%, confidence interval of 5%, and COVID-19 prevalence of 10% to yield an estimated sample size of 2459 participants. Due to low turnout in daily tests conducted, a sample size of 506 participants was included in the study, which was still a representation of the targeted population assuming a test sensitivity of 85%, confidence interval of 5%, a width of 10%, and COVID-19 prevalence of 5% to 10%. To achieve the necessary accuracy on performance estimates, we determine data for negative cases (by NAAT) using a value of 50% for each estimate.

### Data Collection

We used primary and secondary data to determine the cost components of diagnosis and case management of suspected Covid-19 cases. Questionnaires were administered at the facility level and to individuals seeking COVID-19 testing services for cost data. For effectiveness measure, we relied on endpoint data on project-specific reporting forms that included COVID-19 testing registries, laboratory report forms, patient history, case management forms, contact history forms, competency assessments, and Ag-RDT ease of use assessments.

### Model Structure

Figure 1. and 2. depicts the intervention strategies applied. We applied three strategies for the study scenarios. The first strategy involving the use of Ag-RDT followed by a different diagnosis pathway. Under the first scenario Ag-RDT was used as the first-line diagnosis method, followed by the prioritization of negatives for confirmatory testing by NAAT.

**Figure 1:**
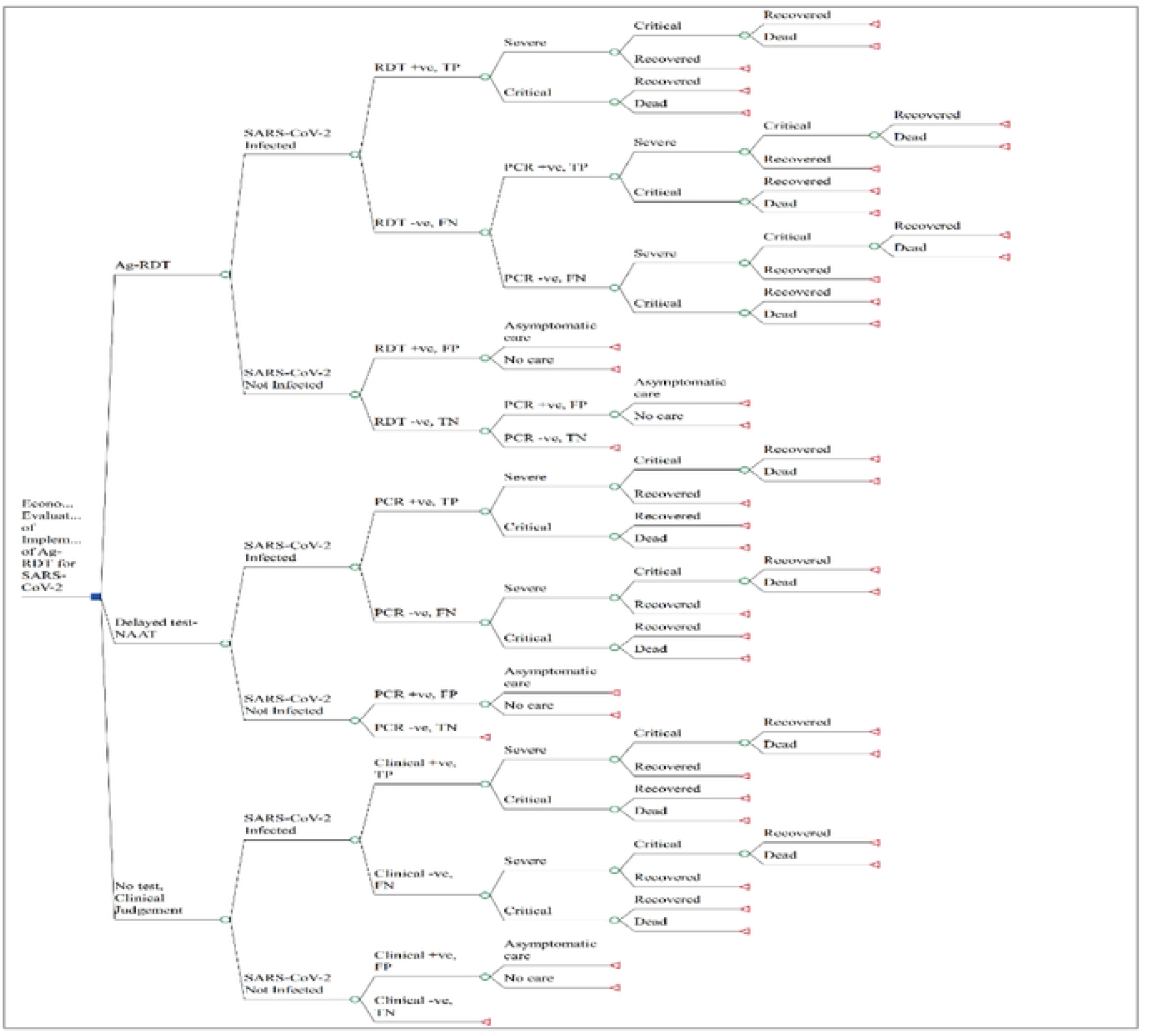
Schematic of decision tree mode under scenario 1. +ve, Positive; -ve, Negative; TP: True Positive; FN: False Negative; FP: False Positive; TN: True Negative

**Figure 2:**
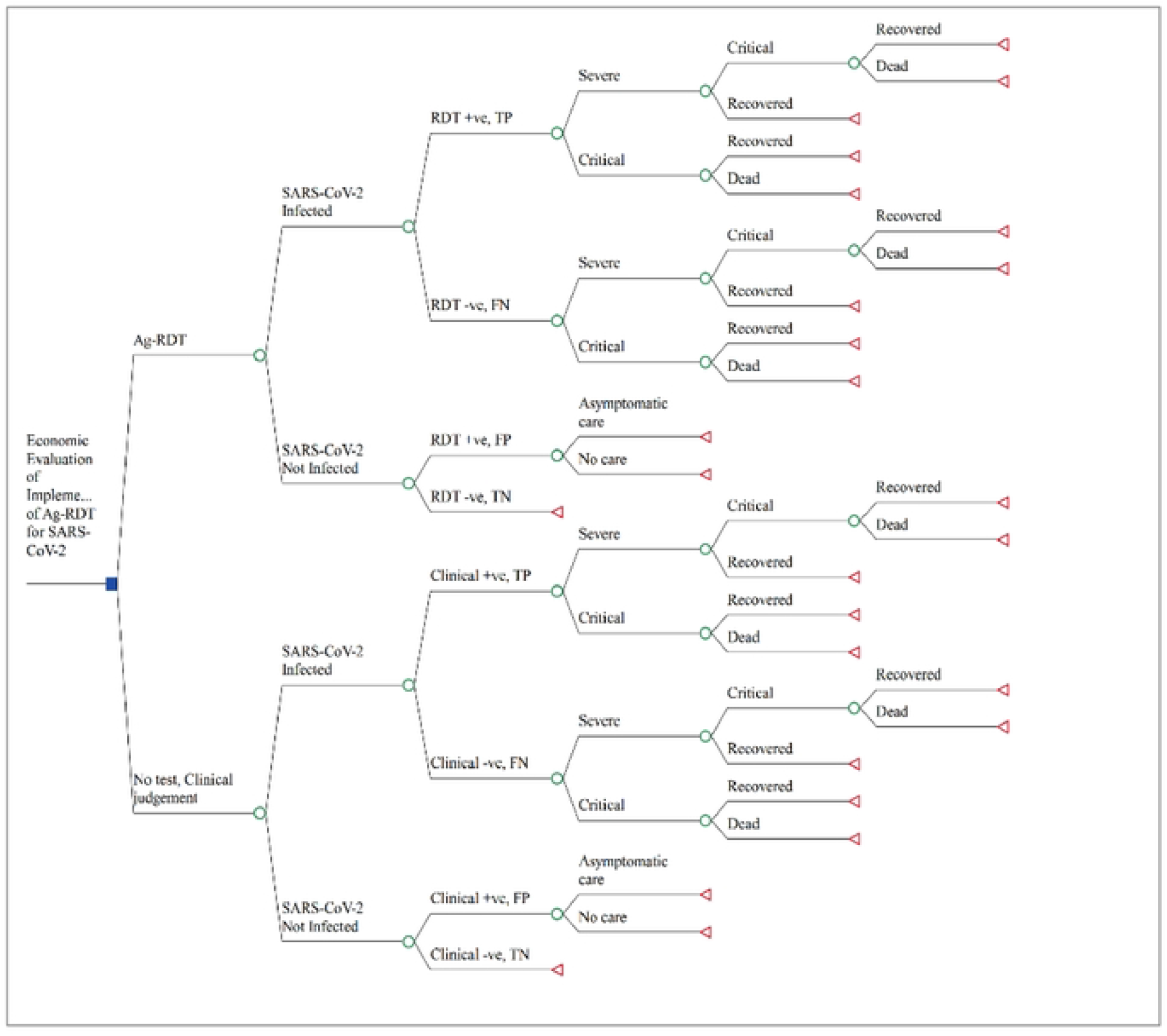
Schematic of decision tree mode under scenario 2. +ve: Positive; -ve: Negative; TP: True Positive; FP: False Positive; FN: False Negative; TN: True Negative

The second scenario involved clinical judgement as the comparator. The first strategy pathway of Ag-RDT diagnosis and case management did not include a confirmatory test of negatives making the diagnosis pathway shorter than in the first scenario where there is access to NAAT services.

### Costing Methods

The costing followed the Global Health Cost Consortium’s (GHCC) reference guidelines [10] to evaluate the cost of implementing SD Biosensor antigen detecting SARS-Cov-2 diagnostic tests in Kenya. We applied an ingredient-based approach from a societal perspective to analyze costs for the diagnosing COVID-19 cases using antigen RDT. Under the healthcare system, we costed both the direct and ancillary costs, which included physical costs and overheads, costs for personal protective equipment (PPE), staff time, and costs for non-pharmaceuticals. We also computed the direct and indirect costs from the patient perspective. For the direct cost, we included the cost of testing, the cost for treatment, the cost incurred for related healthcare services, and the cost of isolation/quarantine. As for the indirect costs, we considered the travel cost; we valued time spent away from normal activities to visit the healthcare facilities; we valued informal care, and using the human capital approach, we valued productivity loss due to absenteeism (figure 3).

**Figure 3.**
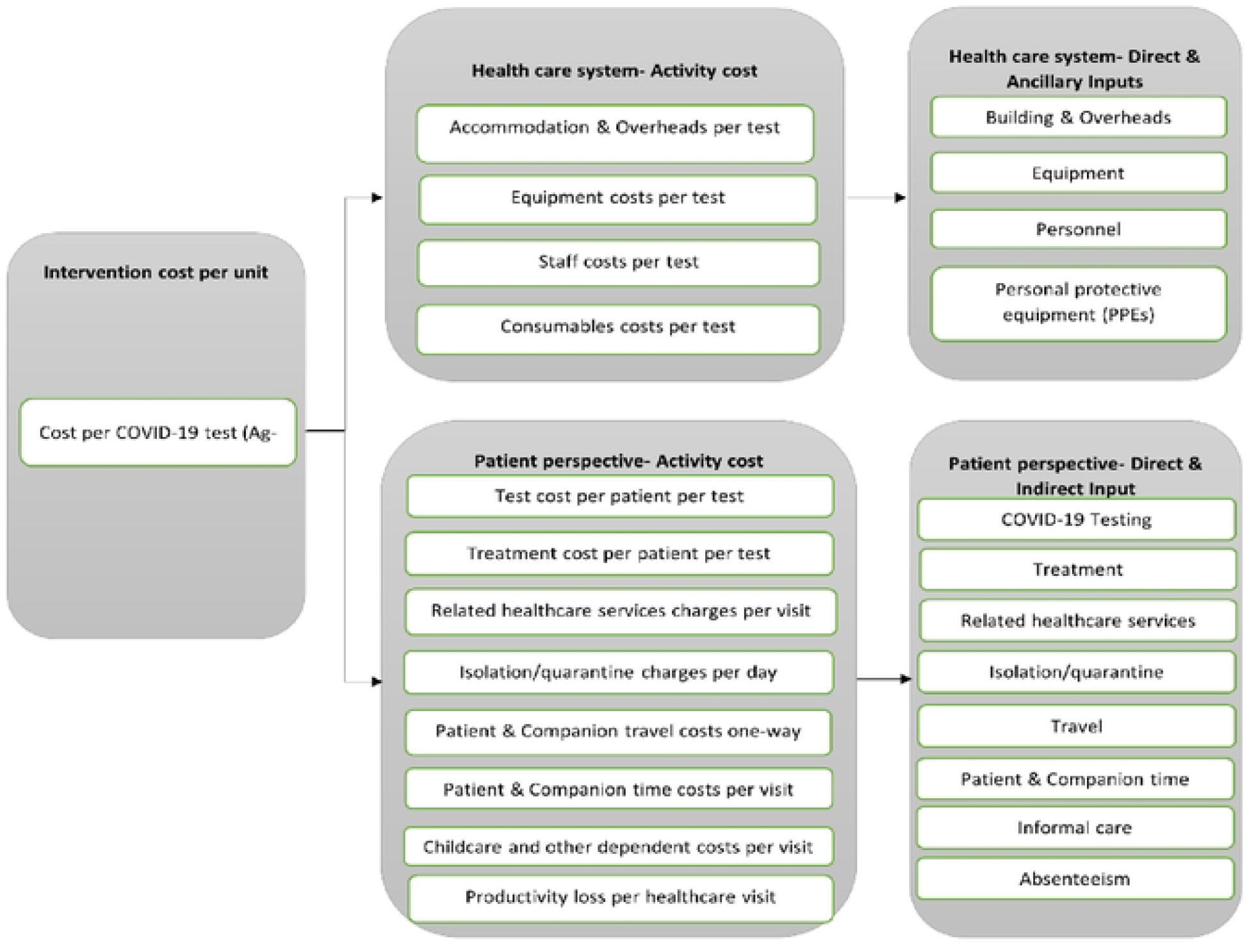
Antigen rapid diagnostic test cost component

The costs for treatment, quarantine, and isolation, such as accommodation and overheads, pharmaceutical, non-pharmaceuticals, staff, PPE, ICU equipment, oxygen therapy, other laboratory tests associated with COVID-19 case management in hospitals, and cost for diagnostic of patients using PCR were derived from a previous study [11].

#### Physical cost and overheads

We obtained outpatient cost overheads from the study on case management of COVID patients [11], which collected primary data from three public health facilities. We computed the physical cost incurred per test by collecting data on the estimated cost of the COVID test room, the size of the facility, and the size of the space the COVID test was being conducted. We later annuitized the estimated cost using the respective useful life years of the housing facility. To estimate the cost of the testing space after annuitizing the cost of the housing facility, we first computed the cost per square metre of the housing facility by dividing the annuitized cost by the size of the housing facility. Second, we multiplied the specific space size for COVID testing by the cost per square metre of the area housing the test. Finally, we divided the cost of the COVID space by the number of tests per day and the number of working days in a year, assuming the daily average test conducted within the last six months and the facility operating every day.

#### Non-pharmaceuticals and Personal Protective Equipment (PPE)

Data was collected on the non-pharmaceuticals and PPE items used during the testing of suspected Covid-19 cases. We obtained cost data for NAAT testing from a recent study by Barasa et.al. (2021) on Examining unit costs for COVID-19 case management in Kenya. The other cost of items for Ag-RDT and quantity required per test were obtained from the sampled health facilities.

#### Staff cost

Data was collected on the type of staff, gross salaries, and time spent on testing from three public health facilities. We computed the amount of time allocated on a test as follows. First, we estimated the total time allocated to testing in a day by obtaining the number of shifts in a day, the number of the specific cadre of staff conducting the test, and the length of each shift in minutes. Second, we estimated the amount of time allocated to a test per day by dividing the number of tests per day, assuming a daily average of tests conducted within the last six months and equal allocation of testing time. Finally, we computed the average staff cost per test by multiplying the staff time allocated to COVID-19 testing in a day in minutes by the gross salary of that cadre of staff per minute.

#### Valuing Time cost

The time patients lost from routine activities was estimated by adding the travel time and the time spent at the health facility as per the patient’s and companion’s response. Using data from Kenya’s minimum wages [12], the time lost was subsequently valued at the average hourly pay of the different categories of paid work the patient and companion would have engaged. While for the unpaid work, a proxy value of the cost of a close market substitute was used.

#### Valuing Productivity Loss

The study considered productivity loss from both paid and replaced unpaid work. Using the human capital approach, the number of hours worked per working day was calculated based on the average number of hours a week the patient worked over the last four weeks, assuming the patient worked for five days in a week. Subsequently, the gross daily wage was estimated by multiplying working hours per day by estimated hourly salaries for different categories of work [12]. Next, the total number of lost productive days from paid work was multiplied by the gross daily wage.

The cost of replacing unpaid work was considered by analyzing the time spent by an informal giver to replace the patient missed unpaid work.

#### Pricing and Valuation

We identified the cost of building as the only capital good, and annuitized it, assuming a useful life of 5 years. We obtained price data from a previous study [11] and presented the costs in Kenya shillings (KES) and US dollars. We used an exchange rate of US$1 = KES 112.52 derived from Xe.com and accessed on 30^th^ November 2021, to convert KES to US$. We obtained shadow prices for unpaid work and the opportunity cost of time from Kenya minimum wages reported by the africanpay.org database accessed on 30^th^ December 2021.

### Effectiveness and Cost-effectiveness measurement

The impact of case management of COVID-19 was dependent on the diagnostic performance of the different diagnostic tests used (Ag-RDT or NAAT), the timing of the test, and the adherence to COVID-19 case management guidelines.

The intermediate outcome was measured in terms of the diagnostic performance of the antigen RDT, which was measured by its sensitivity and specificity compared to the PCR test. Based on the results of 506 test samples, the estimated sensitivity of Ag-RDT is 73%, and the estimated specificity is 93%. Using the diagnostic test confidence interval formula [13], we obtained a 95% confidence interval for the Ag-RDT sensitivity as (59%,87%), and the confidence interval for the specificity as (91%,96%).

The primary health outcome was measured in terms of the cost per disability-adjusted life years (DALYs) averted. We factored in both the mortality and morbidity to obtain DALYs by summing up years of life lost (YLL) and years of life with disability (YLD) [14]. A discounting rate of 3% was used to calculate DALYs and applied Kenya’s life expectancy of 66.34 [15], disability weights as reported [16] and captured in Table.

The incremental cost-effectiveness ratio (ICER) comparing the use of Ag-RDT and confirmatory testing of negatives by NAAT and the use of NAAT as the only diagnostic test conducted was calculated as the difference in costs and DALYs averted of diagnostic and case management in the compared groups.

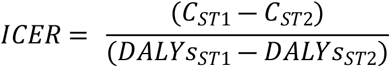

Where ICER = Incremental cost effectiveness ratio; ST1 = RDT as first-line diagnosis followed by NAAT, ST2 = NAAT diagnosis, C_ST1_ = Cost of strategy 1; C_ST2_ = Cost of strategy 2; DALYs_ST1_ = DALYs averted in strategy 1; and DALYs_ST2_ = DALYs averted in strategy 2.

We compared the ICER with an opportunity cost of USD 20.07 to USD 1023.47 (1% to 51% GDP per capita) based on Kenya’s cost-effectiveness threshold as estimated by Woods *et al*. [17] and Ochalek *et al*. [18].

### Assumptions and Parameters

Table 1. present the model parameters. The model also used some assumptions that are key to note. First, we assumed that patients who test positive and show no clinical symptoms of COVID-19 are given home-based standard care, equivalent to isolation and routine care given to mild COVID-19 patients. Second, we assume that false-negative and late diagnosis leads to worsening of symptoms [19]. Third, we relied on a COVID-19 study on an outpatient setting [20] to analyze the outcomes of COVID-19 untreated patients. Lastly, we assumed that all patients who test positive and no further confirmatory diagnostic tests conducted are isolated and provided standard care even though the results could be false positive.

**Table 1:** Key model parameters.

| Description | Value (Lb;Ub) | Source |
| --- | --- | --- |
| **Population** |  |  |
| Study cohort population | 4918 | Author |
| COVID-19 Prevalence | 10% | Author |
| **Cost** |  |  |
| Cost (USD) for asymptomatic care episode | 16.39 | [11] |
| Cost (USD) for conducting a PCR test | 21.84 (21.60;22.87) | [11] |
| Cost (USD) for critical care episode | 472.02 | [11] |
| Cost (USD) for severe care episode | 121.88 | [11] |
| Cost (USD) for conducting a rapid diagnostic test | 4.68 (4.83;7.25) | Author |
| Cost (USD) for clinical diagnosis | 5.22 (5.06;5.36) | Author |
| **DALYs** |  |  |
| **Disability weight for mild COVID-19** | **0.006** | [16] |
| Disability weight for critical COVID-19 | 0.655 | [21] |
| Disability weight for severe COVID-19 | 0.133 | [16] |
| Average age at death | 55.5 | [22] |
| Life expectancy | 66.99 | [15] |
| **Characteristics of Patients** |  |  |
| Proportion of critical patients hospitalized | 0.14 | [23] |
| Proportion of critical COVID-19 who die | 0.892 | [24] |
| Proportion of critical COVID-19 who recover | 0.108 | [24] |
| Proportion of infected patients with SARS-CoV-2 | 0.10 | Author |
| Proportion of patients not given asymptomatic care | 0 | Author |
| Proportion not infected with SARS-CoV-2 | 0.90 | Author |
| Proportion of severe patients hospitalized | 0.86 | [23] |
| Proportion of severe COVID-19 who progress to critical | 0.424 | [25] |
| Proportion of severe COVID-19 who recovered | 0.576 | [25] |
| Proportion of patients given asymptomatic care | 1 | Author |
| Proportion of severe COVID-19 untreated patients who progresses to critical | 1 | Author |
| Proportion of untreated severe COVID-19 patients who recovered | 0 | Author |
| Proportion of untreated critical COVID-19 patients who dies | 1 | Author |
| Proportion of untreated critical COVID-19 patients who recovered | 0 | Author |
| Length of stay asymptomatic care | 12 | [26] |
| Length of stay critical | 7 (4;10) | [24] |
| Length of stay severe | 6 (3;9) | [14] |
| **Diagnostic Accuracy** |  |  |
| PCR Specificity | 0.998 (0.992;0.999) | [27] |
| PCR Sensitivity | 0.86 (0.547;0.994) | [27] |
| RDT Specificity | 0.93 (0.91;0.96) | Author |
| RDT Sensitivity | 0.73 (0.59;0.87) | Author |
| Probability of correctly excluding SARS-CoV-2 | 0.683 (0.60;0.758) | [28] |
| Clinical Judgement Sensitivity | 0.806 (0.729;0.869) | [28] |

### Dealing with uncertainty

We performed a sensitivity analysis on the following parameters: COVID-19 prevalence level; sensitivity of RDT and PCR; the proportion of treated and untreated hospitalized cases; and cost of RDT, NAAT, and treatment of severe and critical cases. We implemented a 20% increase or decrease in the cost of RDT, NAAT, and cost of treatment. The sensitivity analysis of RDT was based on +/- 5% confidence bounds while the bounds of PCR and clinical judgement were provided [27], [28].

We conducted a probabilistic sensitivity analysis to check for the collective uncertainty on the probability of cost-effectiveness using second-order Monte Carlo simulation (n = 1000). We used beta distributions to calculate the probability range of the study parameters and gamma distribution on the cost parameters [29]. Finally, we presented the ICE scatterplot to illustrate the uncertainty in the cost-effectiveness results.

## Results

Table 2. summarizes the key findings from the patient questionnaire administered. As per the results, the primary mode of transport was public transport, with 13 (72%) of the 18 sampled patients preferring public transport to get to the health facility, and the second most popular means was walking on foot, 3 (17%). It was also noted that most (89%) of the patients went alone to the health facility, and only 11% were accompanied. The table also details the patient’s usual activities foregone by visiting a health facility. Most of the patient’s main activities would be, attending to paid work at 28% or attending to a business activity at 28%. Housework activities took 17%, whereas only 6% of patients forego childcare activity.

**Table 2.**
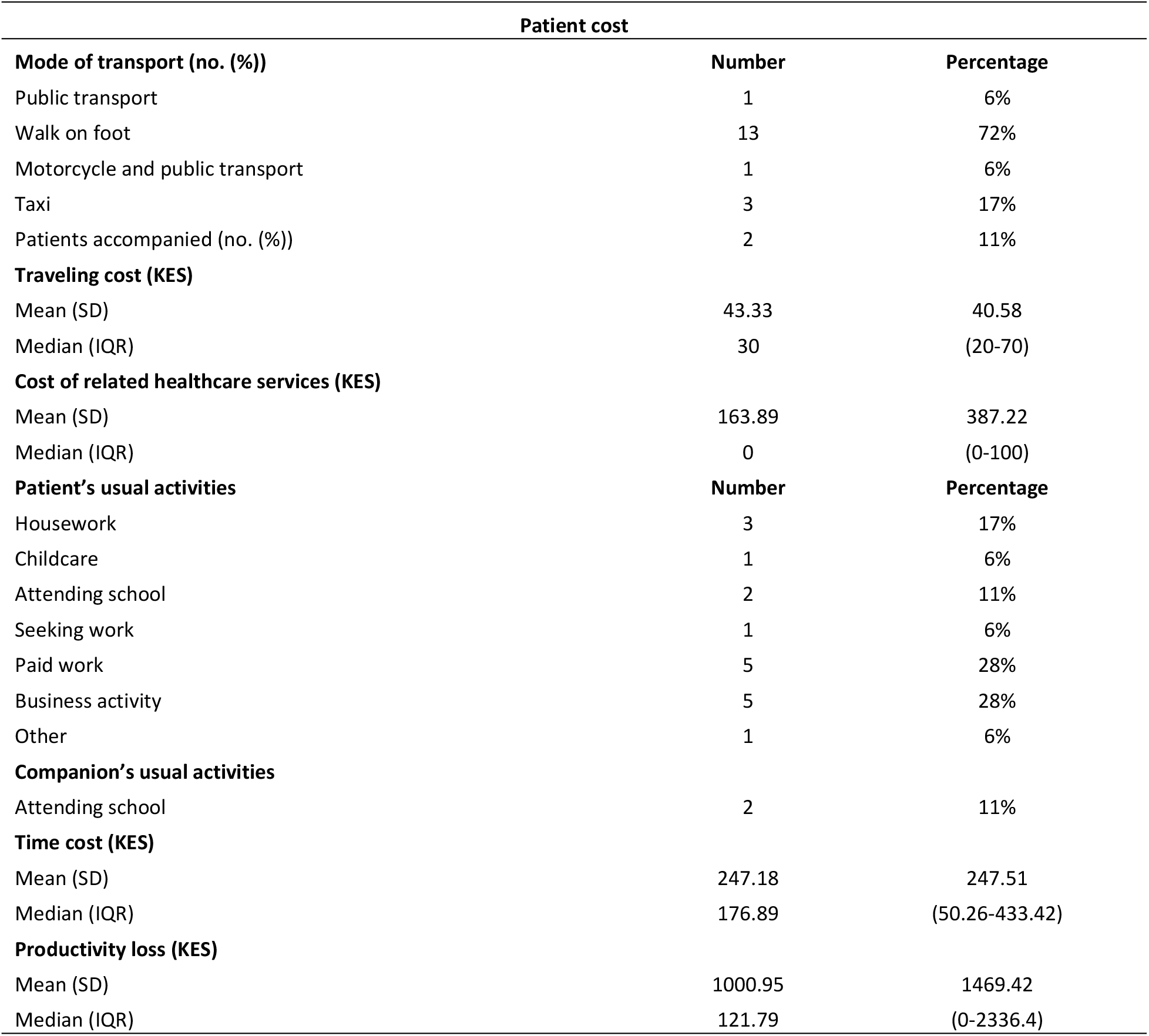
Key findings from the patient’s questionnaire.

| Patient cost |  |  |
| --- | --- | --- |
| Mode of transport (no. (%)) | Number | Percentage |
| Public transport | 1 | 6% |
| Walk on foot | 13 | 72% |
| Motorcycle and public transport | 1 | 6% |
| Taxi | 3 | 17% |
| Patients accompanied (no. (%)) | 2 | 11% |
| **Traveling cost (KES)** |  |  |
| Mean (SD) | 43.33 | 40.58 |
| Median (IQR) | 30 | (20-70) |
| **Cost of related healthcare services (KES)** |  |  |
| Mean (SD) | 163.89 | 387.22 |
| Median (IQR) | 0 | (0-100) |
| **Patient's usual activities** |  |  |
|  | Number | Percentage |
| Housework | 3 | 17% |
| Childcare | 1 | 6% |
| Attending school | 2 | 11% |
| Seeking work | 1 | 6% |
| Paid work | 5 | 28% |
| Business activity | 5 | 28% |
| Other | 1 | 6% |
| **Companion's usual activities** |  |  |
| Attending school | 2 | 11% |
| **Time cost (KES)** |  |  |
| Mean (SD) | 247.18 | 247.51 |
| Median (IQR) | 176.89 | (50.26-433.42) |
| **Productivity loss (KES)** |  |  |
| Mean (SD) | 1000.95 | 1469.42 |
| Median (IQR) | 121.79 | (0-2336.4) |

It also shows the treatment cost, travel cost, time lost per hour, and time cost from the foregone activity the patient would have engaged in during the health facility visit, and the productivity loss. The median travel cost for a one-way visit for a patient was US$0.27. The study findings also report that out of the patients accompanied to the health facility, there was no cost incurred by the patient’s companion while visiting the health facility. Applying the values per hour of paid and unpaid work foregone, the median time cost per hour of both patient’s and companion’s usual activities lost was US$1.57. For the productivity loss, the median productivity cost of absenteeism from both paid and unpaid work was US$1.08.

Table 3. details the unit cost for rapid diagnostic tests compared to NAAT for a COVID-19 suspected case. The results showed the unit cost per test for NAAT and Ag-RDT tests in the healthcare system was US$18.93 and US$3.12, respectively. There is a considerable cost difference between the two tests, mainly because of the laboratory cost incurred when conducting the NAAT test. The table also showed the patient cost incurred for a diagnostic test was US$ 2.92; the major cost driver was the patient time cost. Summarizing the results, we found that the societal cost for PCR was higher at US$21.84 than the Ag-RDT cost of US$4.68.

**Table 3:** Unit costs for antigen RDT and PCR test for SARS-COV-2 detection.

| Inputs | Ag-RDT Test median cost |  | NAAT Test (PCR) cost [11] |  |
| --- | --- | --- | --- | --- |
| | (KES) | US\$ | KES | US\$ |
| **Healthcare perspective** |  |  |  |  |
| Physical cost & overheads per test | 0.23 | 0.00 | 6.66 | 0.06 |
| Equipment per test | - | - | 2.25 | 0.02 |
| Personal protective equipment (PPE) cost per test | 157.72 | 1.40 | 151.76 | 1.35 |
| Consumables cost per test | 31.96 | 0.28 | - | - |
| Lab supplies | - | - | 1959.5 | 17.41 |
| Staff cost per test | 7.57 | 0.07 | 8.98 | 0.09 |
| **Healthcare estimated cost** | **197.49** | **1.76** | **2129.15** | **18.93** |
| **Patient perspective** |  |  |  |  |
| Patient direct cost | 0.00 | 0.00 | 0.00 | 0.00 |
| Patient travel cost | 30.00 | 0.27 | 30.00 | 0.27 |
| Patient time cost | 176.89 | 1.57 | 176.89 | 1.57 |
| Companion travel cost | 0.00 | 0.00 | 0.00 | 0.00 |
| Informal care cost | 0.00 | 0.00 | 0.00 | 0.00 |
| Patient productivity loss | 121.79 | 1.08 | 121.79 | 1.08 |
| **Patient estimated cost** | **328.68** | **2.92** | **328.68** | **2.92** |
| **Societal cost** | **526.17** | **4.68** | **2,457.83** | **21.85** |

### Antigen-RDT and PCR test results

Out of 506 patients recruited, 72 (14.2%) patients tested positive with antigen RDT, 52 (10.3%) patients tested positive with PCR test, 38 (7.5%) were positive for both RDT and PCR test, 34 (6.7%) were positive for RDT and negative for PCR, 14 (2.8%) were positive to PCR and negative to RDT, and 468 (92.5%) were both negative for RDT and PCR test.

### Base case results

The costs, DALYs, and the ICER at 10% COVID-19 prevalence level associated with the three strategies are presented in Table 4. Under the first scenario, where we apply Ag-RDT as the first-line test and prioritization of negatives for the confirmatory test by NAAT in comparison to delaying and conducting NAAT, the findings show no-test strategy is dominated. The results show that the RDT strategy is the costliest, followed by the no-test strategy, and NAAT test strategy was relatively less costly compared to the other two strategies. Although the RDT strategy was costly, it is most effective in averting DALYs to NAAT diagnostic strategy, while failure to conduct a test was less effective to Ag-RDT or NAAT. The results also showed the ICER of Ag-RDT strategy compared to NAAT diagnostic strategy was US$964.63 per DALYs averted, hence a cost-effective strategy when we apply Kenya’s maximum cost-effectiveness threshold of US$1003.4. When we compare the three strategy, the results showed no-test strategy was absolutely dominated, and it would be more efficient to apply the Ag-RDT strategy to case scenarios where there were delay NAAT testing than switching to the no-testing strategy.

**Table 4:** Cost-effectiveness results for Ag-RDT implementation (USD 2021)

| **Scenario 1: Point of care with delayed access to NAAT** |  |  |  |  |  |  |
| --- | --- | --- | --- | --- | --- | --- |
| **Strategy** | **Cost (USD)** | **Incremental Cost (USD)** | **Effectiveness (DALYs)** | **Incremental Effectiveness (DALYs)** | **Incremental cost per DALY averted** | **Dominance** |
| Delayed test-NAAT | 1 107 117.83 (1 102 586.64 to 1 110 014.44) |  | 2236.49 (2229.01 to 2239.81) |  |  | undominated |
| No test, Clinical Judgement | 1 261 230.4 (1 258 276.99 to 1 264 097.03) | 154 112.57 | 2361.35 (2360.40 to 2362.44) | -124.86 | -1234.26 | abs. dominated |
| Ag-RDT | 1 336 231.13 (1 332 224.48 to 1 339 721.30) | 229 113.3 | 1998.97 (1996.95 to 1999.87) | 237.51 | 964.63 | undominated |
| **Scenario 2: Point of care with no access to NAAT** |  |  |  |  |  |  |
| **Strategy** | **Cost (USD)** | **Incremental Cost (USD)** | **Effectiveness (DALYs)** | **Incremental Effectiveness (DALYs)** | **Incremental cost per DALY averted** | **Dominance** |
| Ag-RDT | 998 260.67 (970 948.12 to 1 011 265.58) |  | 2537.8 (2330.70 to 2638.70) |  |  | undominated |
| No test, Clinical judgement | 1 261 230.78 (1 258 276.99 to 1 264 097.03) | 2 62 970.12 | 2361.35 (2330.70 to 2638.70) | 176.45 | 1490.34 | undominated |

Under the second scenario, where Ag-RDT is the only feasible tool to aid testing, the no-test strategy is costly compared to Ag-RDT diagnostic strategy. As for effectiveness, the results show no-test strategy is more effective in averting DALYs than the RDT strategy but with an ICER of US$1490.34 no-test strategy was not cost-effective in Kenya.

### Sensitivity Analysis

#### Difference prevalence level from 5% to 20%

A one-way sensitivity analysis showed the ICER was sensitive to the covid-19 prevalence level. The results (S1 Table) showed that at less than 5% covid-19 prevalence level and under a case where there was access to delayed NAAT, the use of RDT and further confirmatory by NAAT strategy was not cost-effective compared to the delayed NAAT strategy. At a prevalence rate of more than 5% to 20%, the results showed that the use of RDT and further confirmatory of negatives by NAAT was cost-effective compared to the delayed NAAT strategy.

In a scenario with no access to NAAT assay, at a lower prevalence rate of 5% to 16.25%, no-test strategy was still not cost-effective compared to the RDT strategy (S2 Table). The results showed at a higher prevalence rate of 20%, the no-test strategy was more costly and more effective than the Ag-RDT strategy, and the ICER was US$989.15 hence a cost-effective strategy.

#### RDT and PCR Sensitivity

When we varied the sensitivity of RDT (S3 Table) by increasing or reducing RDT sensitivity, we found applying RDT as the first-line tool to aid in testing, followed by prioritization of negatives for confirmatory testing by NAAT was still costly and more effective up to a sensitivity level ≥ 87% to delayed NAAT diagnostic strategy.

In a scenario where there was no access to NAAT assay, RDT was still less costly and less effective than the no-test strategy (S4 Table) and in the two scenarios we found the ICER was sensitive to changes in RDT sensitivity.

When we varied the PCR sensitivity by increasing it, we found PCR was less costly and less effective than RDT. While reducing the PCR sensitivity also led to reduction in the costs of PCR diagnostic strategy and was attractively effective under the three strategies (S5 Table).

According to Figure 4. the key parameters that had the most significant effect on the ICER when we compared the RDT diagnostic strategy to the delayed NAAT diagnostic strategy are 1) Proportion of severe patients hospitalized 2) Proportion of critical patients hospitalized (both of which fewer cases improves cost-effectiveness); 3) Probability of critical patient dying (lower mortality for critical patients improves cost-effectiveness); 4) Length of stay for critical patients (shorter length of stay in the hospital improves cost-effectiveness).

**Figure 4:**
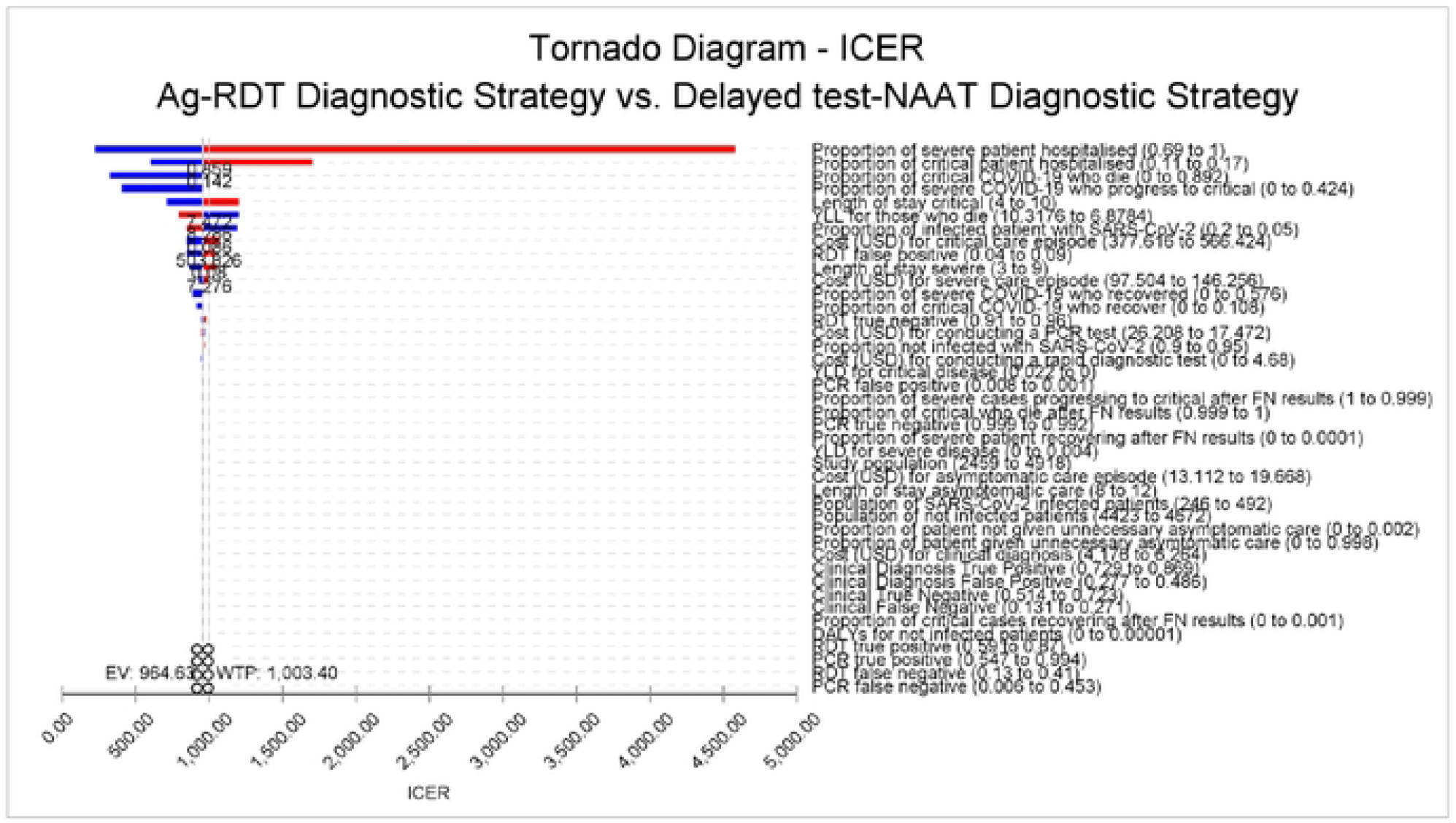
Tornado diagram of one-way sensitivity analysis of the parameters affecting the ICER under scenario 1. YLL, years of life lost; YLD, years of life lived with disability; RDT, rapid diagnostic test; DALYs, disease life adjusted years

Comparing RDT diagnostic strategy and no-test strategy, Figure 5. summarizes the three parameters that had the most significant effect on the ICER. These are: 1) Clinical true positive; (reduction in true positive cases improves cost-effectiveness); 2) Clinical false positive (reduction in false-positive diagnosed cases improves cost-effectiveness, and 3) Proportion of infected SARS-Cov-2 (reduction in SARS-CoV-2 infection improves cost-effectiveness).

**Figure 5:**
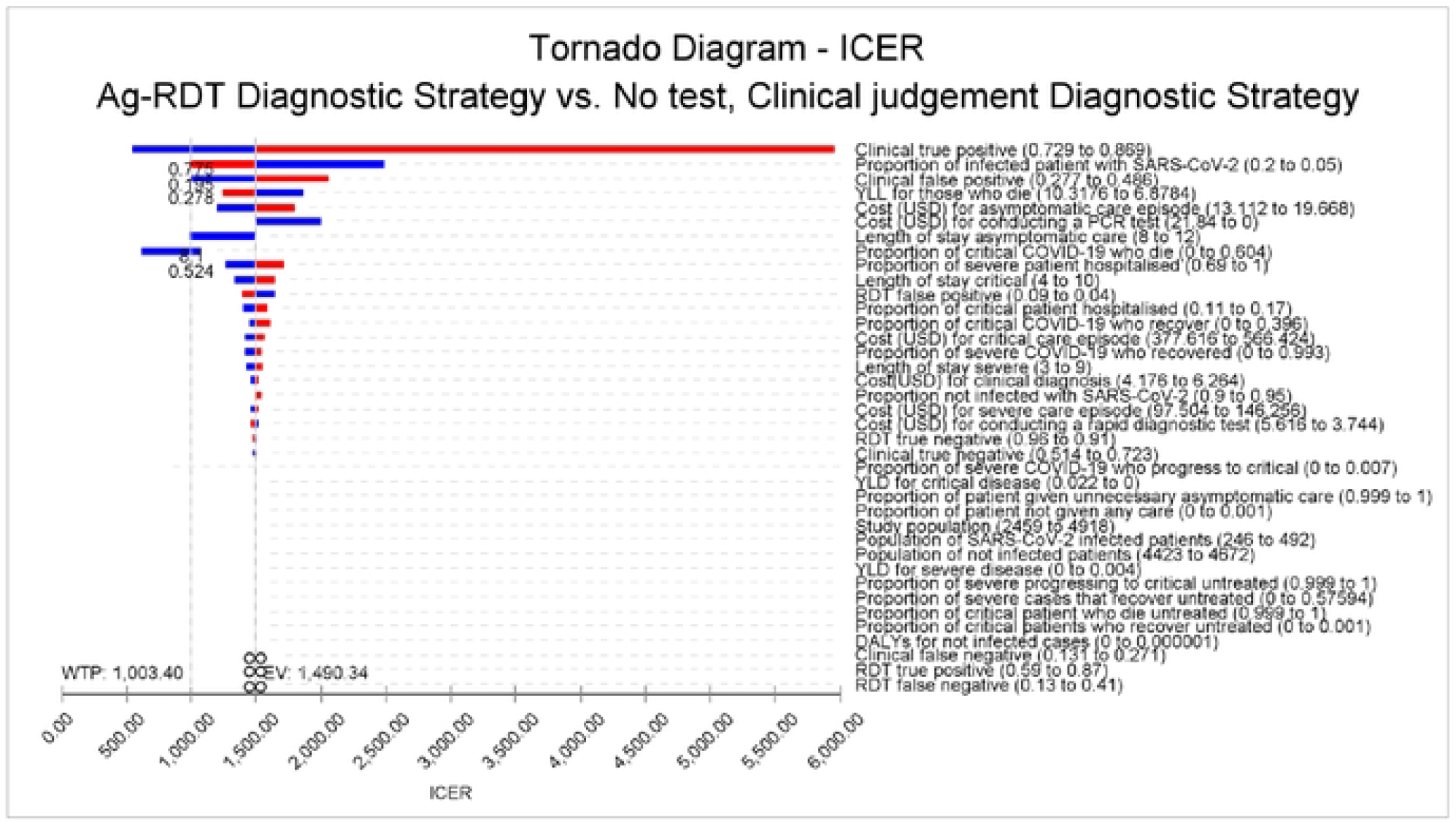
Tornado diagram of one-way sensitivity analysis of the parameters affecting the ICER under scenario 2. YLL, years of life lost; YLD, years of life lived with disability; PCR, polymerase chain reaction; RDT, rapid diagnostic test; SARS-CoV-2, severe acute respiratory syndrome coronavirus 2

#### Probabilistic Sensitivity Analysis

The results of the Monte Carlo simulation of 1000 samples under the first scenario (Figure 6.) show that at a cost-effectiveness threshold of US$ 1003.4 per DALYs averted, the probability of antigen rapid diagnostic test being the more cost-effective strategy was 52.5%. Under the second scenario, the results for PSA (Figure 7.) show that at a cost-effectiveness threshold of US$ 1003.4 per DALYs averted, the probability of the no-test diagnostic strategy being more cost-effective was 28.7%.

**Figure 6:**
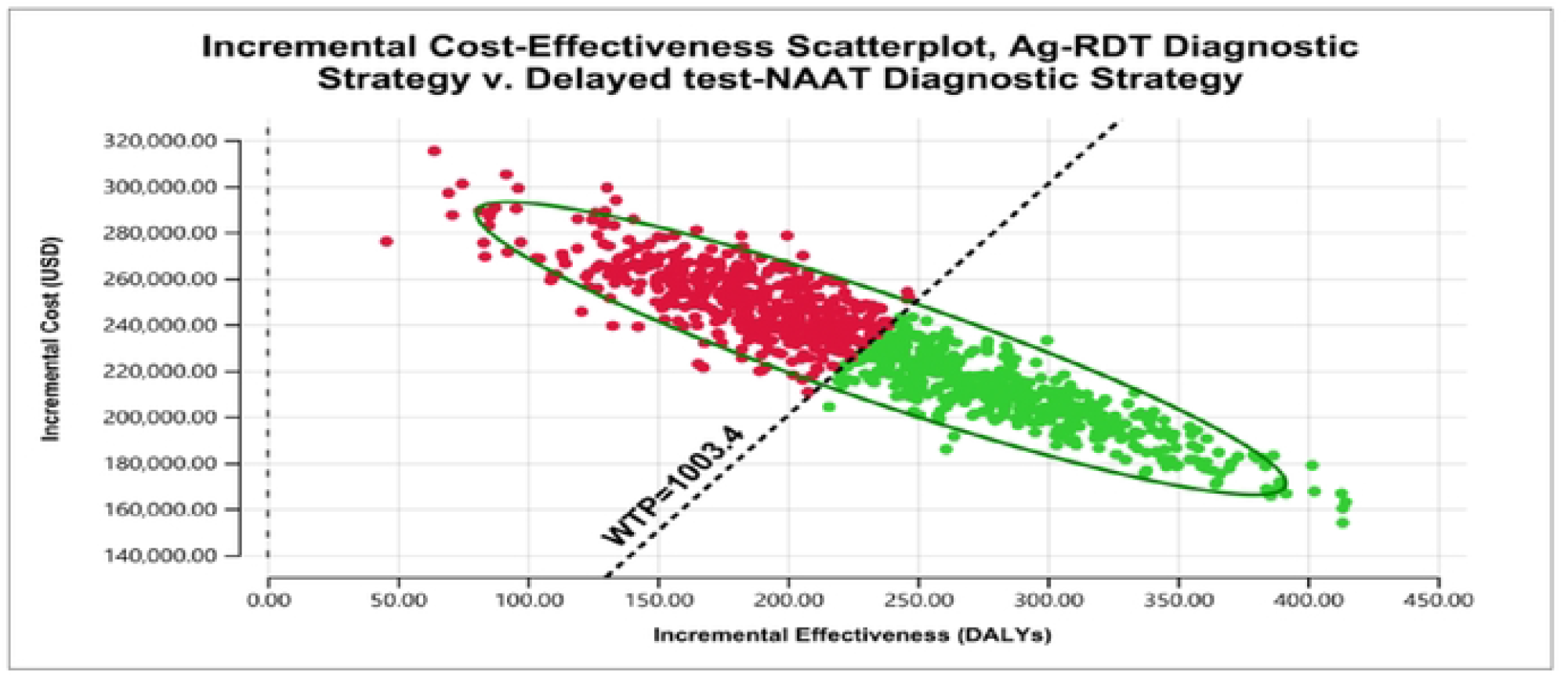
Probabilistic sensitivity analysis of Ag-RDT diagnostic strategy versus delayed nucleic acid amplifying test diagnostic strategy under scenario 1. Green dots representing the points that are cost-effective (below the willingness to pay (WTP)). While the red dots represent the points that are not cost-effective (above the WTP)

**Figure 7:**
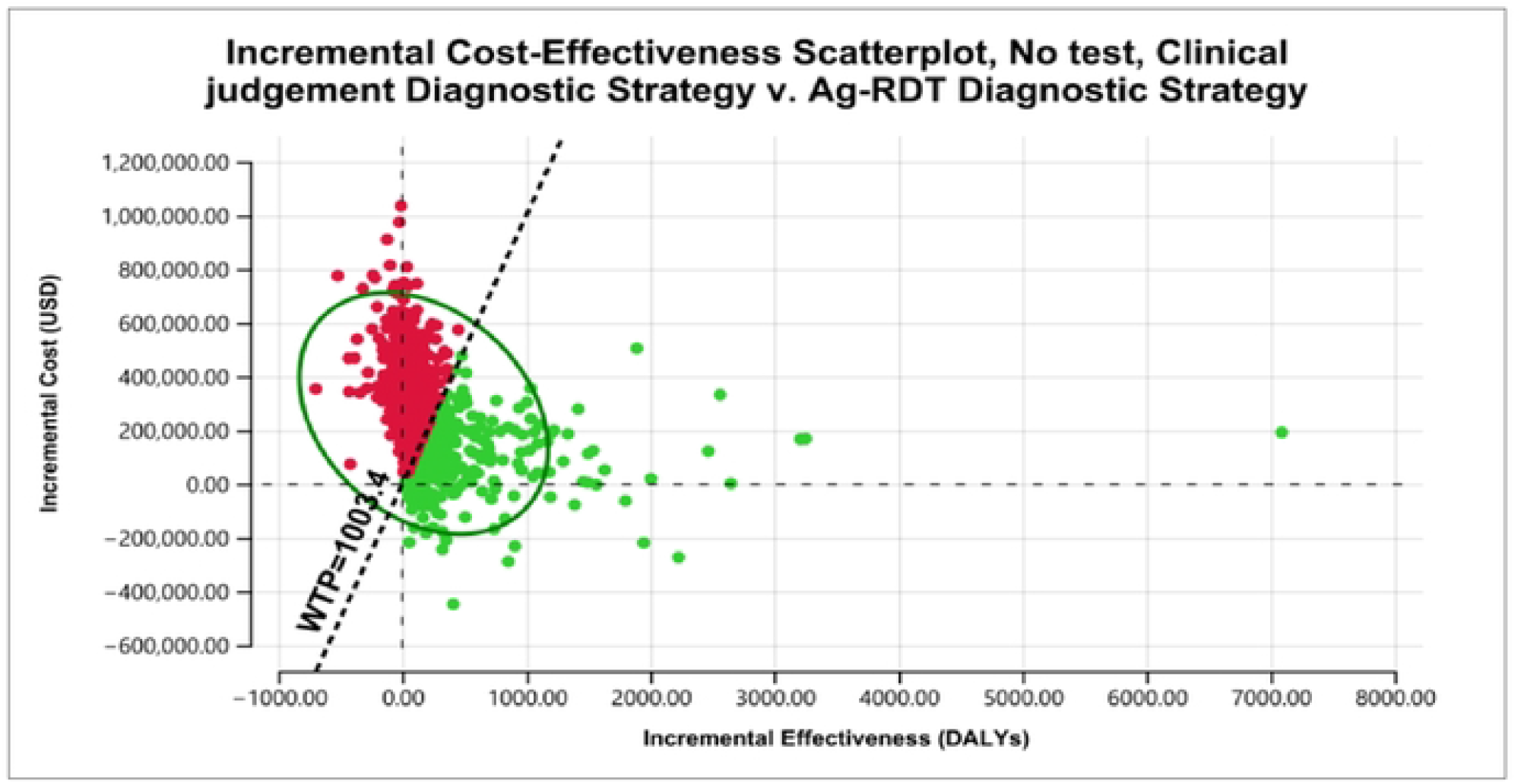
Probabilistic sensitivity analysis of Ag-RDT diagnostic strategy versus delayed nucleic acid amplifying test diagnostic strategy under scenario 1. Green dots representing the points that are cost-effective (below the willingness to pay (WTP)). While the red dots represent the points that are not cost-effective (above the WTP)

Figures 8 and 9 present cost-effectiveness acceptability curves under scenario one and scenario, two respectively based on a range of cost-effectiveness thresholds. Under a scenario where there is delayed NAAT diagnosis and given a willingness to pay of US$ 900 per DALYs averted, there was a 40% probability of the Ag-RDT strategy being cost-effective. The cost-effectiveness acceptability curve shows the probability of the AG-RDT strategy being more cost-effective as the decision maker was willing to increase their willingness to pay (Figure 8). Under a scenario where there is no access to NAAT assay in a resource-limited setting and a decision maker is not willing to pay for any DALYs averted, the probability of Ag-RDT being cost-effective compared to no-test strategy was 94%.

**Figure 8:**
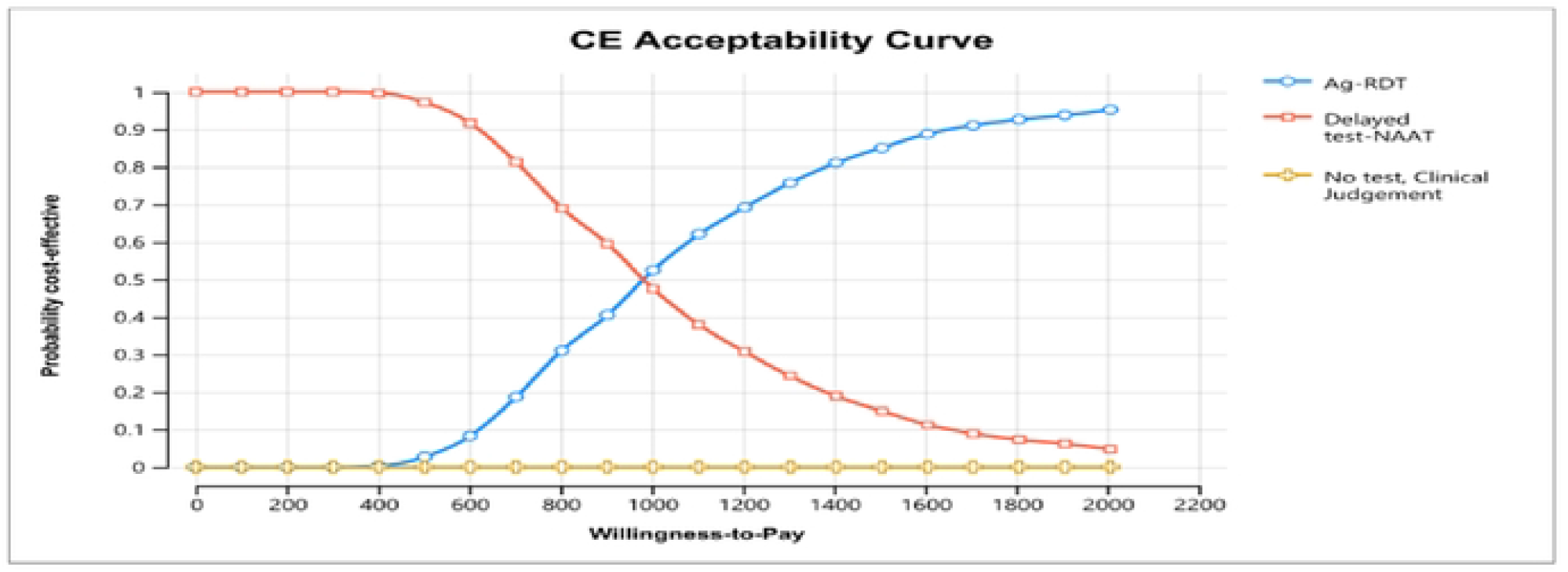
Cost-effectiveness acceptability curve showing the probability of cost-effectiveness of Ag-RDT strategy compared to Delayed test NAAT strategy and No-test Strategy over a range of willingness-to-pay values.

**Figure 9:**
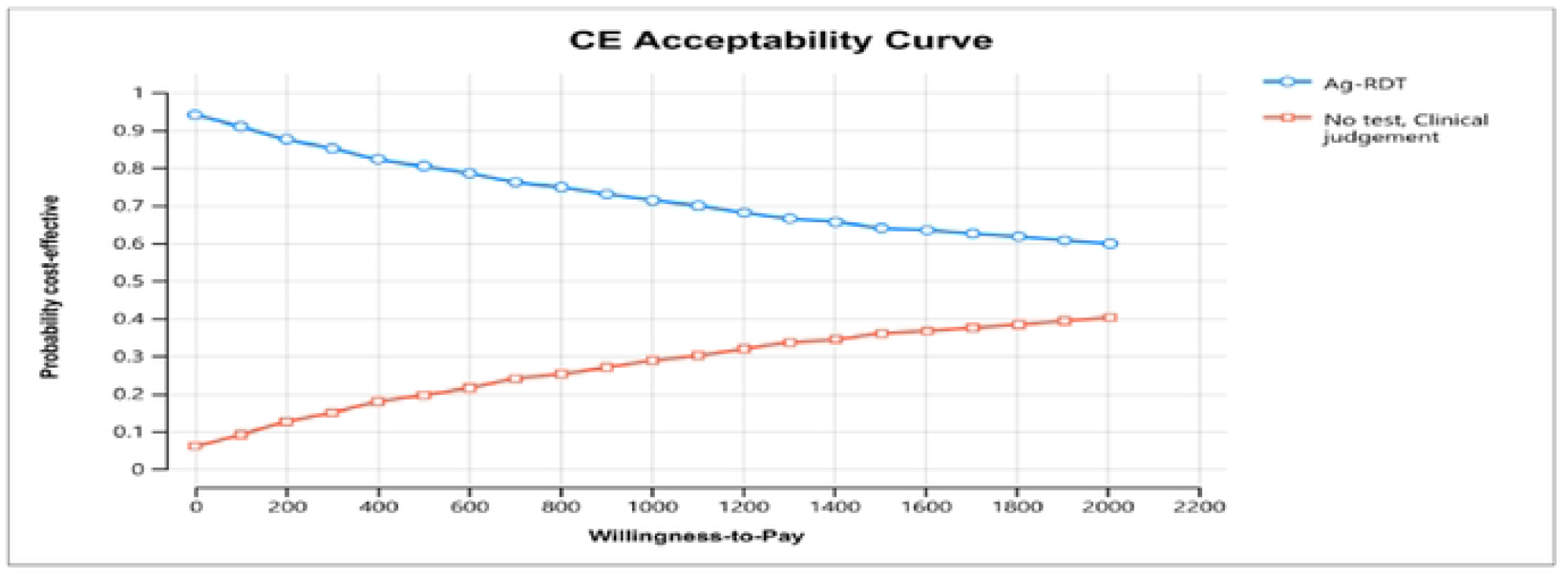
Cost-effectiveness acceptability curve showing the probability of cost-effectiveness of Ag-RDT strategy compared to No-test Strategy over a range of willingness-to-pay values.

## Discussion

The study presents the cost and cost-effectiveness of Ag-RDTs over PCR and clinical judgement for SARS-CoV-2 detection. We compared the use of Ag-RDT under three scenarios using three diagnostic strategies. Our findings show that when we compare the first strategy of using Ag-RDT as the first-line tool and later conducting confirmatory tests of negatives to the second strategy of delaying the testing and using the services of NAAT assays. Ag-RDT diagnostic strategy is costly compared to delayed NAAT diagnostic strategy, the high cost being driven by increased detection of true positives on a confirmatory test of RDT negatives results using PCR test hence increase in the cost of case management of diagnosed positive cases. We also find that using RDT and subjecting the negative RDT results to confirmatory PCR test averted more DALYs on infected SARS-CoV-2 patients not detected by RDT. When we compare the two strategies at ≥8.75% Covid-19 prevalence level, we find that using Ag-RDT as a first-line tool and later conducting confirmatory tests of negatives was a cost-effective strategy.

When we compared the use of Ag-RDT as the only feasible tool to aid in detecting SARS-CoV-2 to the use of clinical judgement, at ≤16.25% Covid-19 prevalence level, the results show no-test strategy was not cost-effective compared to the use of Ag-RDT strategy. The results of the two strategies show that Ag-RDT is less costly and less effective than the clinical strategy, which is substantially more costly and more effective. We can explain the high-cost findings associated with clinical diagnostic strategy to treat presumptive cases with clinical symptoms similar to SARS-CoV-2 infected patients. The clinical diagnostic strategy averts more DALYs than the RDT strategy since most cases with SARS CoV-2 clinical symptoms are subjected to care/treatment. Still, in a resource-limited setting, in the case of Kenya, the strategy may not be cost-effective due to the high cost associated with the strategy. At a higher prevalence level, a presumptive diagnosis of SARS-CoV-2 has been found to have a higher sensitivity level and a relatively lower specificity level [30]. The results show at a higher Covid-19 prevalence level (20% prevalence), the use of a no-test, clinical judgement strategy was cost-effective in averting DALYs compared to the use of Ag-RDT strategy at a relatively high willingness to pay threshold.

Our findings show the proportion of severe and critical cases hospitalized impacts more on the cost-effectiveness of the Ag-RDT strategy. This could be explained by the fact that a low (high) proportion of severe and critical cases hospitalized implies a low (high) Covid-19 prevalence level, which in turn provides a reason for an accurate diagnosis to avert more DALYs and cost implications associated with misdiagnosis. When we consider delayed NAAT strategy previous studies have shown late diagnosis may not be associated with ICU admission or death [19] hence delay in obtaining results or access to NAAT cannot be linked to the patient disease progression and recovery. Although, it can be argued that late diagnosis of COVID-19 patients can increase the risk of infection, especially contact with individuals such as caregivers, as personal protective equipment may not be used. When considerations are made to implement RDT diagnostic strategy in settings with delayed access to NAAT, it will avert more DALYs than delay NAAT diagnostic strategy.

The major limitation of this study is the scarce data on the outcomes of COVID-19 patients with false-negative diagnosis results. However, we made assumptions based on disease progression and outcome during the peak of the COVID-19 pandemic for the COVID-19 cases that were not given any care. One strength of this analysis is the fact that it considered the diagnostic cost and the treatment cost associated with false-positive cases. This paper is among the first papers on cost effectiveness of Ag RDTs in low- and middle-income countries.

## Conclusion

The study findings should inform policymakers to support the implementation of the Ag-RDT diagnostic strategy in a scenario where there is delayed access to confirmatory NAAT testing. At a high prevalence level, the use of Ag-RDT diagnostic kits would be a cost-effective strategy compared to delaying and applying the NAAT testing strategy. Under a scenario where there is no access to NAAT assay, the optimal strategy would be to support the use of RDT rather than resorting to clinical judgement as a strategy for diagnosis. Since the use of Ag-RDT would be a cost-saving strategy and an optimal strategy in a resource-limited setting like Kenya. There is an increased opportunity for cost-effectiveness and cost savings if Ag-RDT is introduced to complement the use of NAAT assay where there are delays in confirmatory testing and scenarios where there is no access to NAAT assay. The implementation and roll-out of Ag-RDT will reduce the risk of misdiagnosis and case management of false positive cases, especially in a rural setting where due to lack of NAAT assay there may be an overreliance on clinical judgement to diagnose Covid-19 suspected cases.

## Data Availability

PLOS Data Policies

https://kemriwellcometrust-my.sharepoint.com/:x:/g/personal/barwah_kemri-wellcome_org/EcxX2qi3po5Hlj_la-GT5oUBC4aCpKvfYLnJIQa224bT-w?e=6PWOaZ

## Ethical considerations

This research was approved by the Ethics committee of Kenya Methodist University and all participants signed a written consent form to participate to the study

## Policy implication

This paper will give important insight on cost effective tests to use in a pandemic and will help decision makers to use efficiently the scarce healthcare resources

## Funding

The project that generated data used in this study was made possible by the generous support of the World Health Organization. The study was an implementation Research on the use of Antigen Rapid Diagnostic Tests for Coronavirus Disease 2019 (COVID-19). The study entailed the assessment of field performance, feasibility, acceptability, ease of use and impact of Ag-RDTs for the diagnosis of SARS-CoV-2 infection in Kenya. The funding was awarded to JG. The funders had no role in study design, data collection and analysis, decision to publish, or preparation of the manuscript.

## Acknowledgement

We are grateful to the Mount Kenya University study team fieldworkers who collected effectiveness data.

## Supporting information

**S1 Table:** Different prevalence levels from 5% to 20% sensitivity report scenario 1

**S2 Table:** Different prevalence levels from 5% to 20% sensitivity report scenario 2

**S3 Table:** RDT Sensitivity Report Scenario 1

**S4 Table:** RDT Sensitivity Report Scenario 2

**S5 Table:** PCR Sensitivity Report

## Notes

### Competing Interest Statement

The authors have declared no competing interest.

### Clinical Protocols

https://kemriwellcometrust-my.sharepoint.com/:w:/g/personal/barwah_kemri-wellcome_org/EbhHYk8oaW5Gm1-IC5PuaQwBzkpLJTy3vOb5D2AIvHhmTg?e=HLBzfF

